# Epidemiological, clinical Characteristics and mortality of patients Infected with SARS-CoV-2 Admitted of Kinshasa University Hospital, Democratic Republic of the Congo from March 24, 2020 to January 30, 2021: Two waves, two faces?

**DOI:** 10.1101/2021.09.05.21262678

**Authors:** Madone Mandina, Jean-Robert Makulo, Roger Wumba, Ben Bepouka, Jerome Odio, Aliocha Nkodila, Murielle Longokolo, Nadine Mayasi, Donatien Mangala, Guyguy Kamwiziku, Auguy Luzayadio Longo, Guillaume Mpia, Yamin Kokusa, Hervé Keke, Marcel Mbula, Hippolyte Situakibanza, Ernest Sumaili, Jean-Marie Kayembe

**Affiliations:** Kinshasa University Hospital, Faculty of Medicine, University of Kinshasa, DRC; Unit of Vaccinology, World Health Organization, Kinshasa, Democratic Republic of the Congo; Technical Secretariat of the Multisectoral Committee for the Response to Covid-19, DRC

**Keywords:** SARS-COV-2, Democratic Republic of Congo, 2 waves

## Abstract

**Background:** The objective of our retrospective study was to establish a comparison between the first and the second waves of demographic and clinical characteristics as well as mortality and its determinants.

**Methods:** A total of 411 COVID-19 patients were enrolled in Kinshasa University Hospital and categorized into two groups according to the pandemic pattern, demographics, and disease severity. The clinical characteristics were compared according to the two waves. To describe survival from the first day of hospitalization until death, we used Kaplan Meier’s method. We used the Log Rank test to compare the survival curves between the two waves. The Cox regression was used to identify independent predictors of mortality.

**Results:** During the study period, 411 patients with confirmed COVID-19 were admitted to the hospital. The average age of patients in the 2nd wave was higher than in the first wave (52.4 ±17.5 vs 58.1 ±15.7, p=0.026). The mean saturation was lower in the first wave than in the second. The death rate of patients in the first wave was higher than in the second wave (p=0.009). Survival was reduced in the first wave compared to the second wave. In the first wave, age over 60 years, respiratory distress, law oxygen saturation (≤89%) and severe stage of COVID-19 emerged as factors associated with death, while in the second wave it was mainly respiratory distress, law oxygen saturation (≤ 89%) and severe stage. The predictors of mortality present in both the first and second waves were respiratory distress and severe COVID-19 stage.

**Conclusion:** Mortality decreased in the second wave. Age no longer emerged as a factor in mortality in the second wave. Health system strengthening and outreach to those at high risk of mortality should continue to maintain and improve gains.

## Introduction

Beginning in December 2019 in China, the coronavirus disease 2019 (COVID-19) caused by the severe acute respiratory syndrome coronavirus 2 (SARS-CoV-2) has spread rapidly around the world and was declared a pandemic on 11 March 2020 ^1^. By the end of January 2021, there had been 103 500 950 confirmed cases worldwide, including 2 240 771 deaths, with a lethality rate of 2.2% ^2^. Africa was largely spared by the pandemic initially, unlike the countries of the West. However, since December, the Covid-19 seems to have hit harder in a hitherto relatively untouched Africa. Indeed, the number of infected people and the number of deaths has increased since the beginning of December 2020. At the end of January, there were now officially a total of 3.3 million infected people on the continent, 700,000 more than three weeks ago, according to data from Johns Hopkins University. However, the number of infections and mortality is still much lower than in Europe or the United States ^2,3^.

The Democratic Republic of Congo (DRC) recorded its first confirmed case on March 10, 2020, and, undoubtedly, by a simple coincidence, the day after the declaration of this first case, the disease was officially declared a “pandemic” by the WHO ^1^. Following the first declared cases, a state of health emergency was declared on 24 March 2020, with the closure of national and international borders, schools and universities, churches, markets, etc. ^4^. For obvious reasons of economic constraints linked to poor countries, total containment as applied in the West could not be recommended ^5^. Did these early measures protect the country from a more serious pandemic? At the end of January 2021, there were 23 043 confirmed cases in the DRC, including 18 206 cases (78.2%) in the city province of Kinshasa, the epicenter of the disease, with a total of 674 deaths, a lethality of 2.9% ^6^. However, a significant drop in Covid-19 cases has begun to be observed across the country since July 2020. Several Covid-19 treatment centers (COTC) in the capital have even had to close their doors due to a lack of confirmed cases. This situation justified the lifting of the state of health emergency with a phased resumption of activities ^7^. In November 2020, an upward trend in cases was observed in the following months, confirming a second wave of the pandemic linked to Covid-19, similar to that observed in Europe ^8^.

Among the COTCs in the city province of Kinshasa, the COTC of the Kinshasa University Hospital opened in March 2020, which, in the meantime, has seen its technical facilities improve considerably with better prepared staff. The question is recurrent, and rather legitimate: what is the difference between these two waves? As the context and the strategies implemented at the country and COTC levels are not the same, comparing the two waves could yield interesting information. The objective of our retrospective study is to establish a comparison between the first and the second waves of demographic and clinical characteristics as well as mortality and its determinants.

## Methods

### Study design and period

We studied a series corresponding to all cases of Covid-19 hospitalized in a treatment center in the city of Kinshasa, the epicenter of Covid-19 in the Democratic Republic of Congo between March 24, 2020 and January 31, 2021. All patients admitted up to June 30 were considered to be in the first wave and all those admitted on or after July 1 were in the second wave. Every wave lasted three and a half months.

The study was conducted in a large covid-19 treatment center (COTC) located at Kinshasa University Hospital, a hospital attached to the country’s largest university, the University of Kinshasa (UNIKIN). The COVID-19 center has a capacity of 50 beds with a permanent staff specialized in Intensive Care and Infectious Diseases, and has a medical biology laboratory capable of diagnosis by RT-PCR. The COVID-19 center can also provide emergency hemodialysis, interventional endoscopies, and chemotherapy to COVID-19 cancer patients.

### Study population

the study population included all patients hospitalized at the COTC during the study period. The criteria for hospitalization of patients were: to have a positive Covid-19 test with a moderate to severe or critical form evaluated according to World Health Organization (WHO) interim guideline ^9^, a very suggestive clinical context with a pathological CT scan in the case of a negative test. All cases of benign Covid-19 were treated at home.

### Inclusion criteria

this study included all patients with a covid-19 confirmed by a positive test. The cases with a negative covid-19 test were not included.

Information related to demographic data, clinical characteristics, treatment and outcomes of patients was collected retrospectively.

### Operational definitions

Any patient with clinical indications and/or visual signs on chest CT that are suggestive of Covid-19 is a suspected case of Covid-19.

Any symptomatic patient with RT-PCR and/or IgM or IgG positive is a confirmed case.

The criteria for hospital discharge: were defined by the absence of fever for at least 3 days, clinical remission of symptoms and/or a negative RT-PCR test on the 12th day of hospitalization (10).

### Statistical analysis

statistical analysis was performed with SPSS Statistics Software (version 21; IBM, New York, USA).

Data is given as numbers and percentages or means and standard deviations. Statistical comparisons between two groups were made using the χ2 test (categorical variables) or the student’s t test. Kaplan Meier’s method described survival from the first day of hospitalization until death (complete data) to the end of the study (censored data). We used the Log Rank test to compare the survival curves between the two waves. The Cox regression was used to identify independent predictors of mortality. Statistical significance was set at p < 0.05.

### Ethical considerations

The study was approved by the University of Kinshasa School of Public Health’s Ethics Committee (ESP/CE/179/2020). The data was collected anonymously and confidentially. The privacy and personality of the patients were safeguarded. Because of the retrospective nature of the study and the minimal risk posed by it, obtaining informed consent was not considered necessary.

## Results

### General characteristics of the study population

During the study period, 411 patients with confirmed COVID-19 were admitted to the hospital. The number of patients admitted was 215 in the first wave and 196 in the second wave. The average age of patients in the 2nd wave was higher than in the first wave (52.4 ±17.5 vs 58.1 ±15.7, p=0.026). The age group over 60 years was the most common among patients and was more prevalent in the second wave than in the first. Most patients come directly from the house. The number of patients coming from home in the first wave was higher than in the second wave. Heart disease was more common in the first wave than in the second.

Patients in the first wave differed from those in the second wave in that they had a higher frequency of headaches, fever, coughs, and throat pain. The average saturation was lower in the first wave than in the second. Patients with very low oxygen saturation (<50%) were more frequently encountered in the first wave. Signs of respiratory distress were more common in the first wave than in the second.

Corticosteroids, anticoagulants, and antibiotics were prescribed more in the second wave than in the first. Patients with anemia and high urea levels were more frequently encountered in the first wave (Table 1).

**TABLE 1.** The WHO clinical classification of the Covid-19 (9)

| Mild case | Moderate | Severe | Critically severe |
| --- | --- | --- | --- |
| Mild clinical manifestations, no imaging performed | Fever, dyspnea, Appearance of pneumonia on Xray or CT | Meet any of the following:<br>Respiratory distress,<br>RR ≥ 30/min<br>Oxygen saturation ≤ 93%<br>PaO<sub>2</sub>/FiO<sub>2</sub> ≤ 300 mmHg | Respiratory failure requiring enhanced respiratory support, which may be combined with shock or other organ dysfunction, patients need intensive care monitoring and treatment |

**Table 1.**
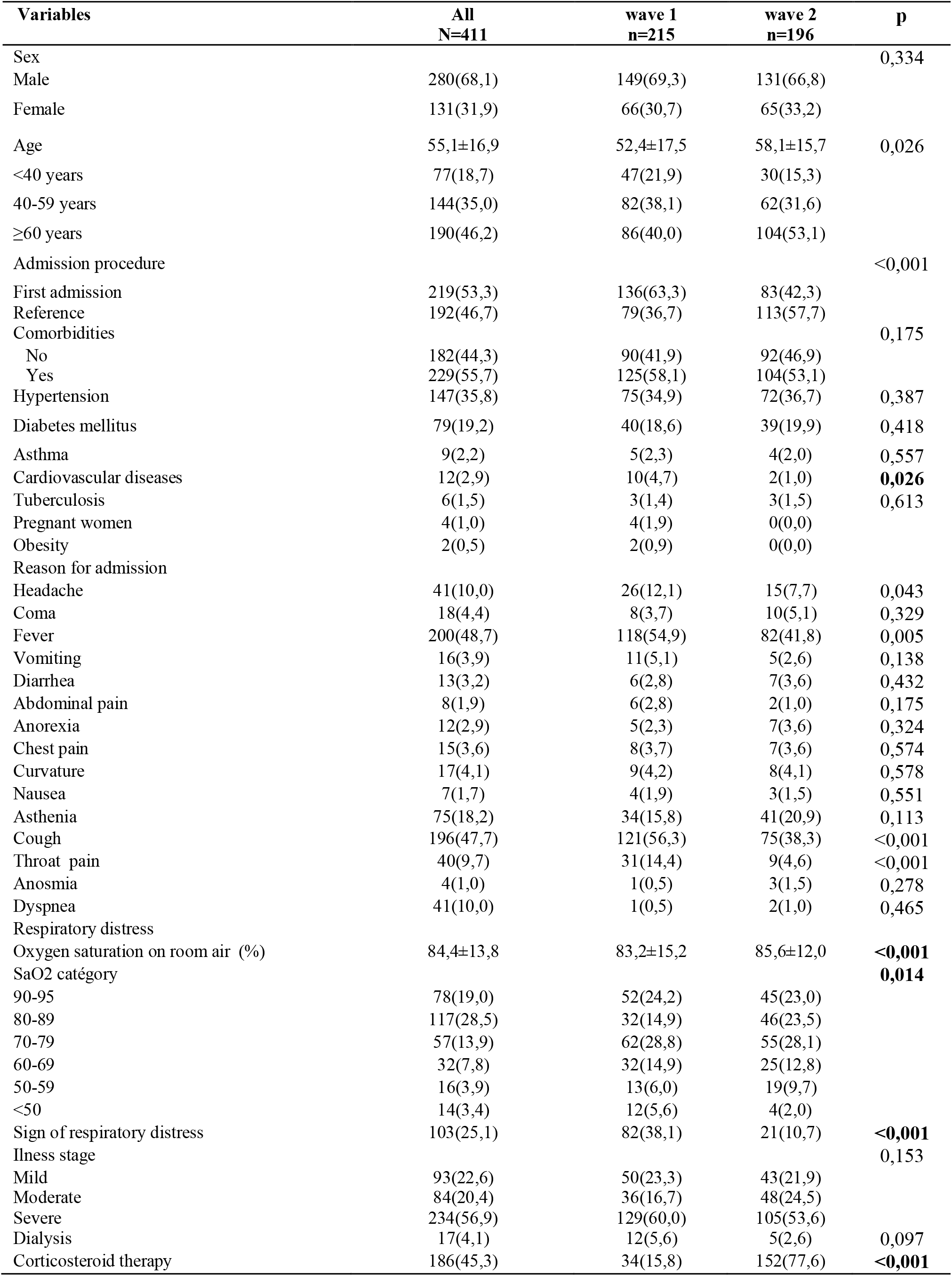
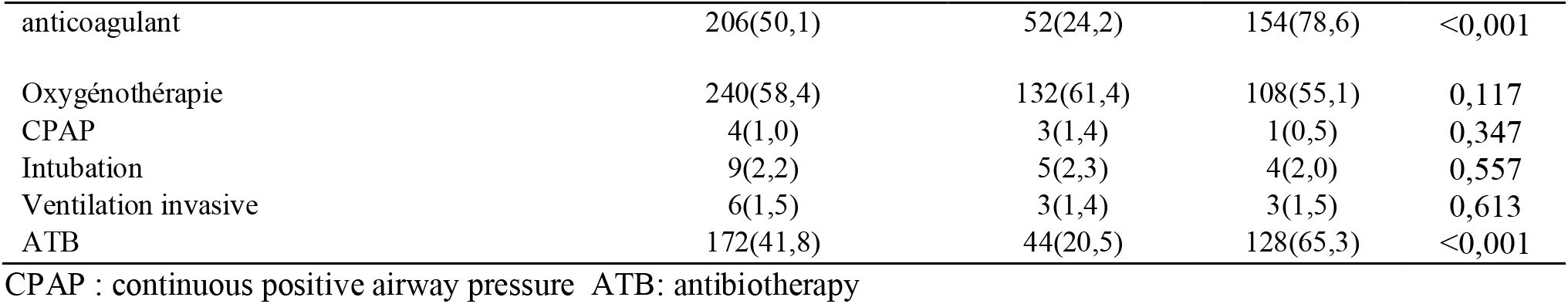
General characteristics of 411 patients admitted to the COVID-19 treatment center of the Kinshasa University Hospital between March 24, 2020 and January 31, 2021

### Vital outcome of patients by wave

Death of patients in the first wave was higher than patients in the second wave (p=0.009) (Figure 1).

**Figure 1.**
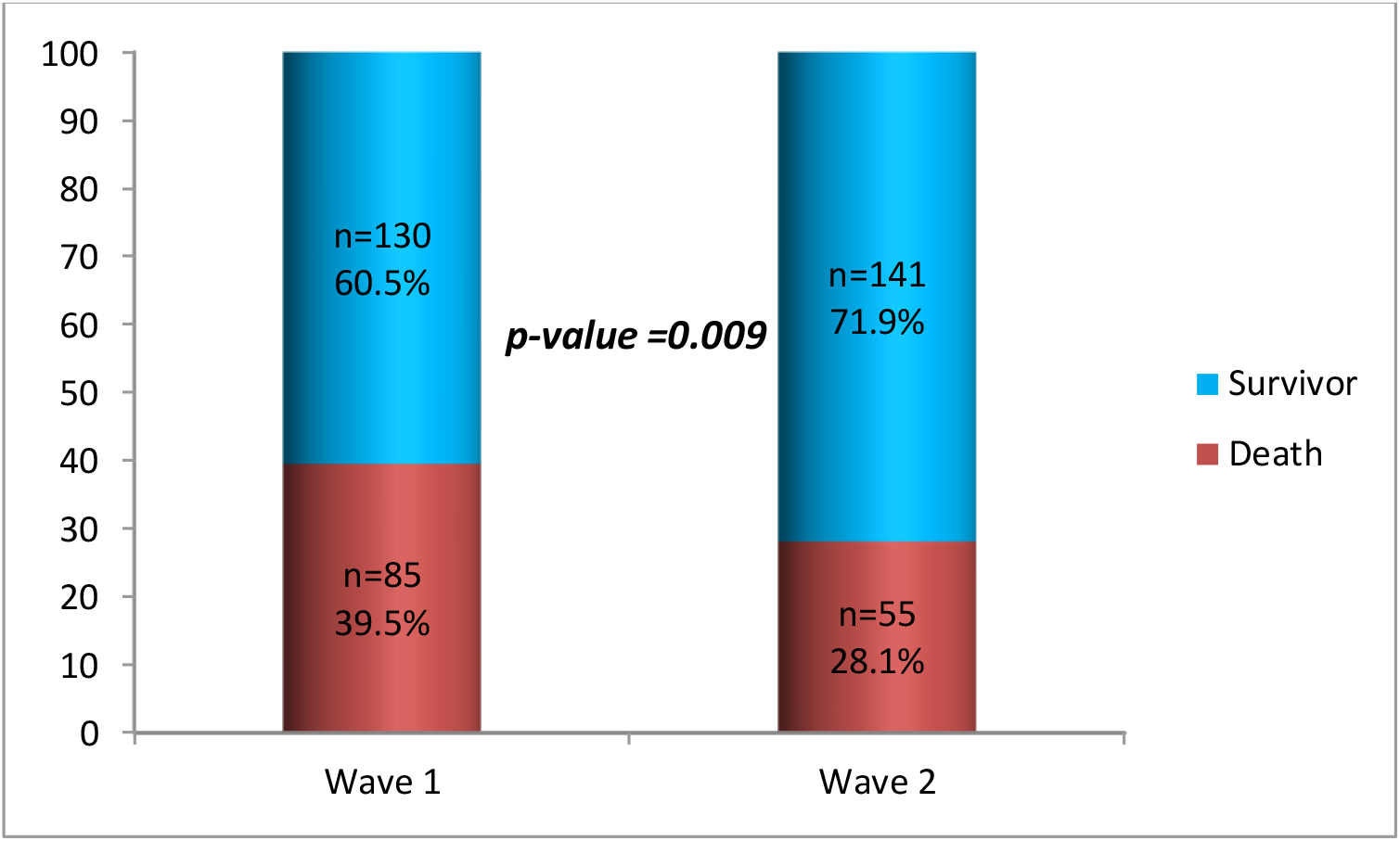
Vital outcome of patients by wave.

### Survival of COVID-19 patients

#### Overall survival of both waves

**Figure 2.**
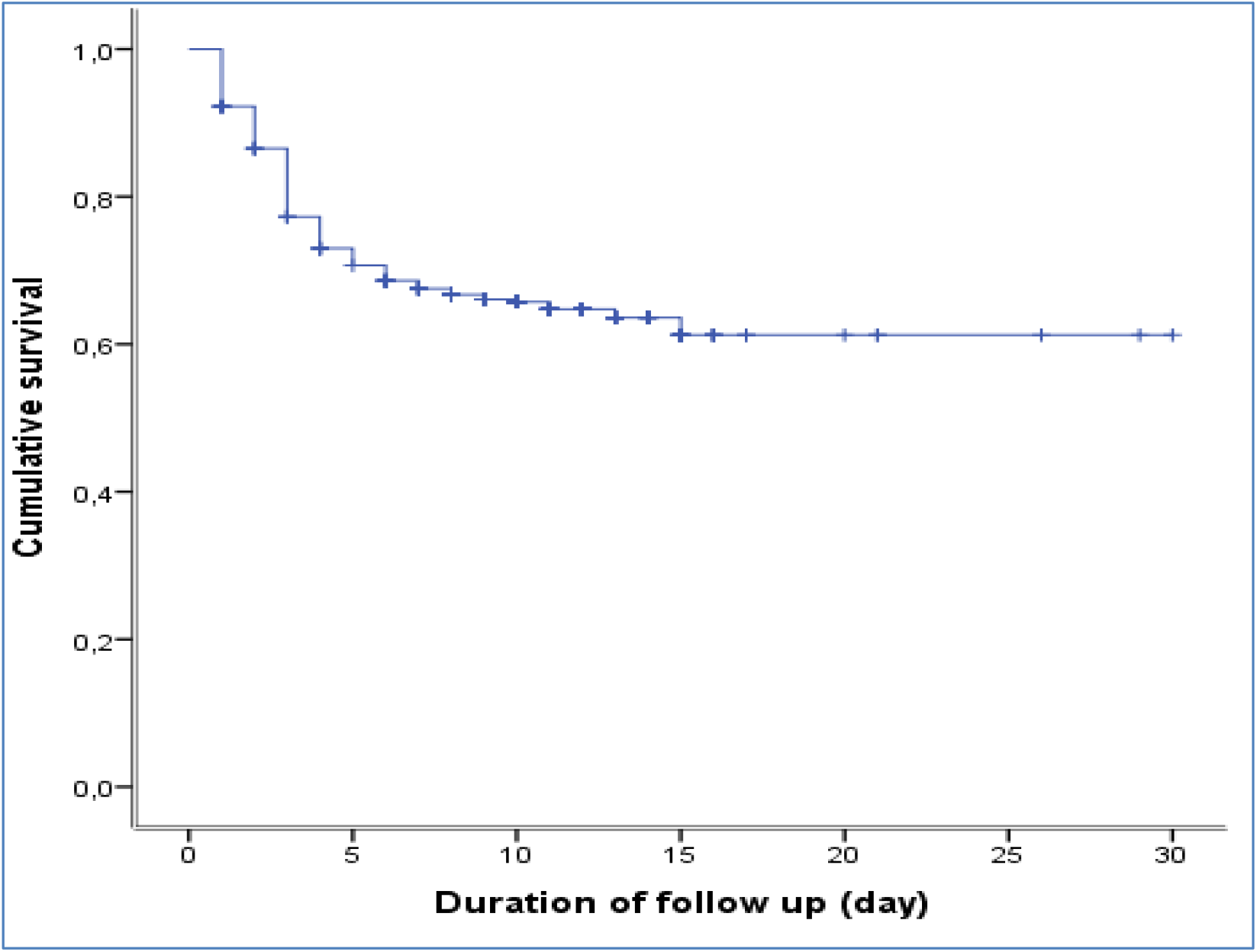
Overall survival of both waves.

Day 0=100%

Day 1=92,2%

Day 3=77,6%

Day 5=71,3%

Day 10=66,9%

Day 20=65,9%

Median overall survival is 9 (8-10) in living 10 (8-11), deceased 3 (1-4)

#### Survival according to wave

Survival was reduced in the first wave compared to the second wave. At 15 days of follow-up, about 40% of the patients in the first wave had died, while in the second wave it was about 30% (figure 3).

**Figure 3.**
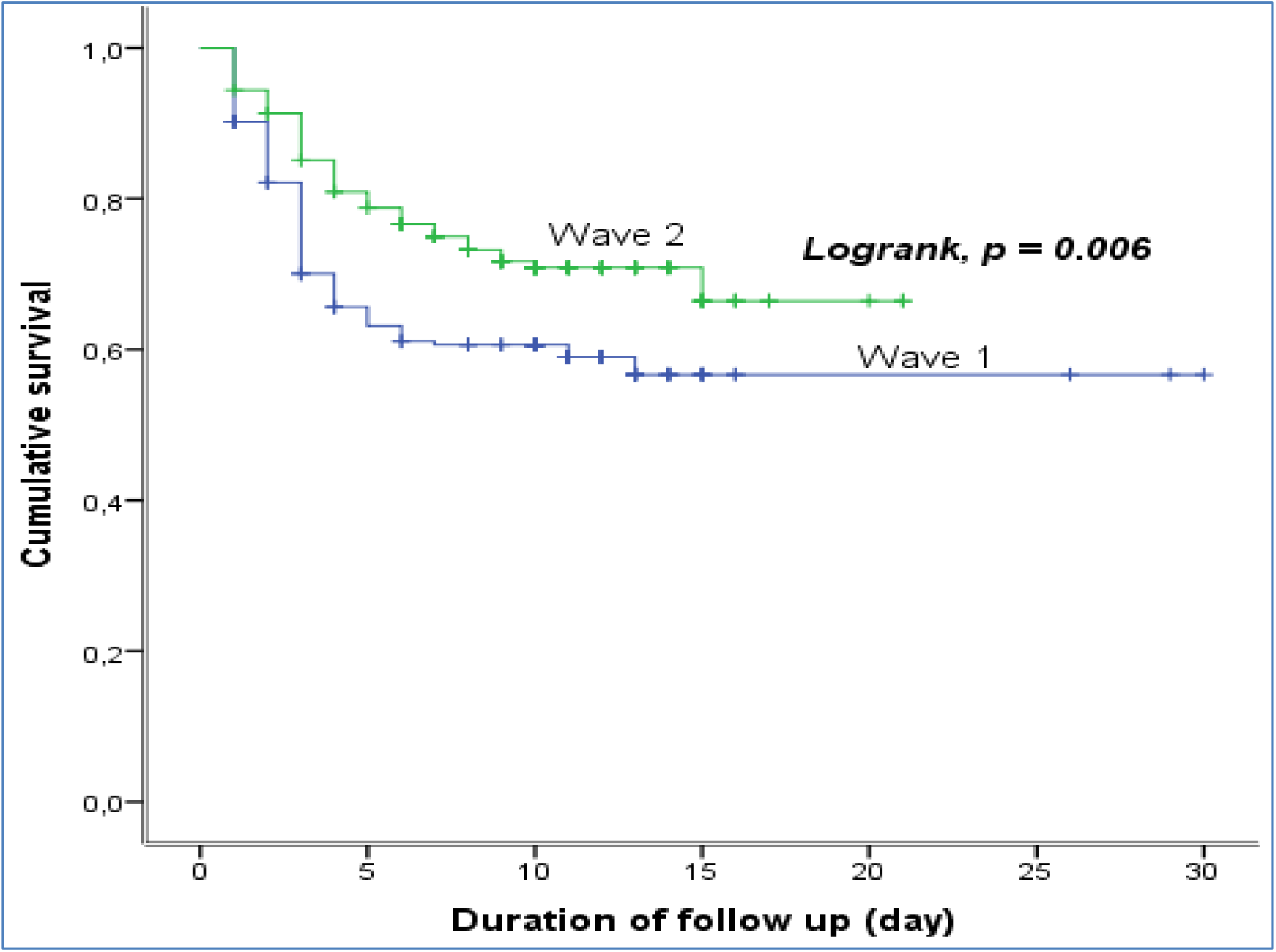
Survival according to waves.

#### Independent Predictors of Mortality in COVID-19 Patients

The predictors of mortality in all patients were age greater than or equal to 60 years, presence of coma, low oxygen saturation (≤89%), and severe stage of COVID-19. In the first wave, age over 60 years, respiratory distress, law oxygen saturation (≤89%) and severe COVID-19 stage emerged as factors associated with death, whereas in the second wave it was mainly respiratory distress, desaturation (≤89%) and severe stage. The predictors of mortality present in both the first and second waves were respiratory distress and severe COVID-19 stage (table 2).

**Table 2.**
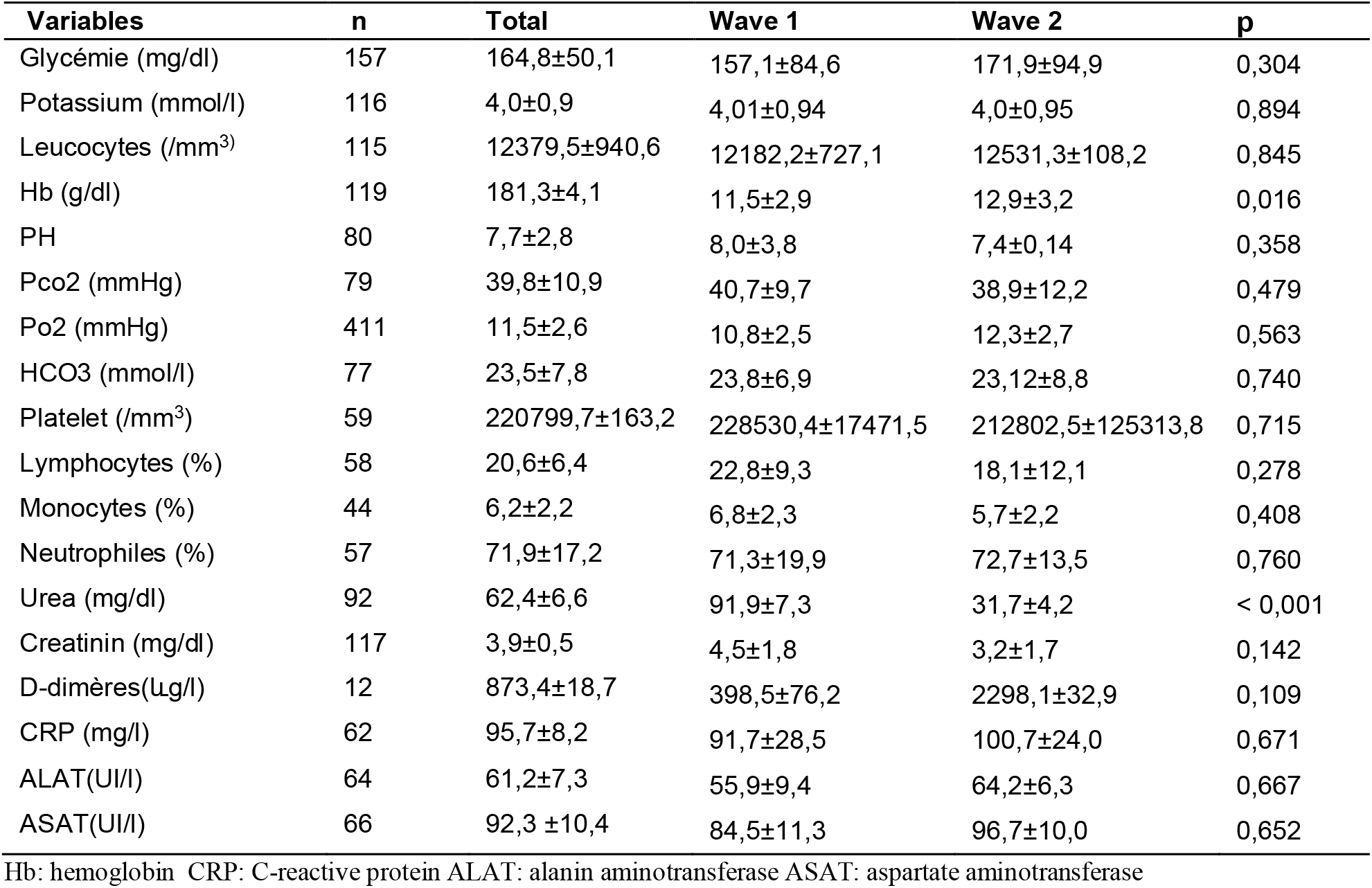
Biologic characteristics of 411 patients admitted to the COVID-19 treatment center of the Kinshasa University Hospital between March 24, 2020 and January 31, 2021

**Table 2.** Independent predictors of mortality in COVID-19 patients admitted to the COVID-19 treatment center of the Kinshasa University Hospital between March 24, 2020 and January 31, 2021

| Variables | All |  | Wave 1 |  | Wave 2 |  |
| --- | --- | --- | --- | --- | --- | --- |
|  | HRa (IC95%) | p | HRa (IC95%) | p | HRa (IC95%) | p |
| Age |  |  |  |  |  |  |
| <40 years | 1 |  | 1 |  | 1 |  |
| 40-59 years | 0,96(0,54-1,72) | 0,902 | 1,25(0,63-2,51) | 0,525 | 0,95(0,31-2,91) | 0,932 |
| ≥60 years | 1,89(1,21-2,74) | **0,016** | 2,02(1,06-3,87) | **0,034** | 1,33(0,47-3,75) | 0,592 |
| Admission procedure |  |  |  |  |  |  |
| First admission | 1 |  | 1 |  | 1 |  |
| Reference | 0,97(0,68-1,38) | 0,865 | 1,19(0,65-2,17) | 0,579 | 1,66(0,85-3,21) | 0,135 |
| Comorbidities |  |  |  |  |  |  |
| No | 1 |  | 1 |  | 1 |  |
| Yes | 1,00(0,69-1,44) | 0,998 | 1,03(0,56-1,91) | 0,915 | 1,07(0,57-2,01) | 0,839 |
| Respiratory distress |  |  |  |  |  |  |
| No | 1 |  | 1 |  | 1 |  |
| Yes | 1,12(0,57-1,65) | 0,572 | 2,50(1,17-5,33) | **0,018** | 2,89(1,39-4,20) | **0,005** |
| Coma |  |  |  |  |  |  |
| No | 1 |  | 1 |  |  |  |
| Yes | 1,99(1,03-3,84) | **0,040** | 1,96(0,78-4,91) | 0,150 | 2,50(1,91-6,88) | **0,016** |
| Oxygen saturation |  |  |  |  |  |  |
| >95 % | 1 |  | 1 |  | 1 |  |
| 90-95 % | 1,27(0,06-1,42) | 0,124 | 2,63(0,86-8,05) | 0,090 | 1,14(0,9-1,41) | 0,919 |
| 80-89 % | 1,53(1,03-4,76) | **0,033** | 2,03(1,04-3,09) | **<0,001** | 1,25(0,63-2,48) | 0,884 |
| 70-79 % | 2,91(1,52-5,56) | **0,022** | 2,45(1,72-5,38) | **<0,001** | 1,32(0,42-2,47) | 0,781 |
| 60-69 % | 3,89(1,92-7,85) | **0,008** | 2,88(1,30-9,58) | **<0,001** | 1,37(0,44-3,09) | 0,419 |
| 50-59 % | 4,52(2,17-9,40) | **<0,001** | 3,9(2,05-9,99) | **<0,001** | 1,42(0,50-3,48) | 0,358 |
| <50 % | 8,50(3,89-18,56) | **<0,001** | 4,6(2,96-13,17) | **<0,001** | 1,66(0,15-8,31) | 0,275 |
| Severity of the disease |  |  |  |  |  |  |
| Mild | 1 |  | 1 |  | 1 |  |
| Moderate | 3,16(0,76-5,07) | 0,113 | 14,13(3,38-59,17) | **<0,001** | 1,83(0,42-6,66) | 0,905 |
| Severe | 4,42(1,87-8,55) | **0,004** | 24,30(7,44-79,42) | **<0,001** | 12,08(1,30-22,56) | **0,029** |

## Discussion

Compared to the 2 waves of COVID-19 in the present study, the average age was higher in the second wave, while comorbidities including heart disease and clinical signs (fever, headache, cough, sore throat, respiratory distress) were more common in the first wave. The essential therapeutic approaches in COVID-19, including corticosteroid therapy and anticoagulation, were used more in the second wave. The mortality rate was higher in the first wave. The factors predicting mortality that emerged in both waves were respiratory distress and severe stage of COVID-19.

In contrast to what has been reported elsewhere; in the present study, patients were older in the second wave. Elsewhere, patients admitted in the second wave were younger. There were fewer deaths in the second wave, in agreement with the results reported by previous research in several countries ^11-13^. The present study did not find differences in the frequency of concomitant diseases between the two waves, results similar to those of other studies ^14^. Although in our study, heart disease was more frequent in the first wave, this was probably due to the awareness of patients with comorbidities to respecting in a particular way the barrier measures. The clinical signs most encountered in the first wave: fever, headache, cough, throat pain, respiratory distress. There were more severe cases in the first wave than in the second. Changes in social policy due to COVID-19, such as changes in the admission criteria for patients with mild symptoms, strongly influence the characteristics of patients who are admitted to our treatment center. In the first wave, all mild cases were admitted, while in the second wave, mild cases were admitted only if they had comorbidities, taking into account the criteria for frailty.

Treatment approaches were strengthened in the second wave. Most patients in the second wave were on corticosteroids and anticoagulants. We noted that early in the coronavirus pandemic, deaths due to COVID-19 were partly attributed to blood clot formation that led to more serious thrombotic events such as pulmonary failure, heart attack, and stroke. These were quite common in patients with severe cases of COVID-19. The paper published in the British Medical Journal shows that patients who receive preventive doses of anticoagulants within the first 24 hours of hospitalization for COVID-19 have a 30% lower mortality rate than patients who do not receive this drug. A study by Zhang et al. showed higher mortality in COVID-19 patients with thromboembolism ^15^. Another study by Tang et al. found significantly higher d-dimer levels at admission in the non-survivor group, indicating a worse prognosis in patients with new-onset coronavirus pneumonia with coagulopathy ^16^. A meta-analysis by Malas et al. reported an overall arterial thromboembolism (ATE) rate of 2%, a venous thromboembolism rate of 21%, a venous thrombosis rate of 20%, and a pulmonary embolism rate of 13% in those infected with SARS-COV-2. They also reported that the odds of mortality were significantly increased by thromboembolism (up to 74%). In contrast, a study by Hippensteel et al. found no significant difference in mortality among critically ill patients, although they did find a higher prevalence of venous thromboembolism in critically ill patients with COVID-19 ^17^. Corticosteroids were used more in the second wave than in the first. The first published randomized trial is a preliminary analysis of the ongoing RECOVERY trial in the UK. This is a randomized, controlled, open-label trial that randomized hospitalized patients with clinically suspected or laboratory-confirmed SARS-CoV-2 infection between usual care without corticosteroids and usual care with dexamethasone. Dexamethasone was administered orally or intravenously at a dose of 6 mg/day for up to 10 days, or until hospital discharge. Patients requiring invasive mechanical ventilation (IMV) and receiving dexamethasone had a reduced death rate than those receiving standard treatment alone (29.3% vs. 41.4%, rate ratio ¼ 0.64: CI95%: 0.51-0.81), as well as in those receiving supplemental oxygen without IMV (23.3% vs. 26.2%, rate ratio ¼ 0.82, CI 95%: 0.72-0.94) ^18^. Based on these convincing results, the same dosage of dexamethasone was routinely used in our patients with severe COVID-19 in the second wave, which may also be a factor for improved management and low mortality in our treatment center.

The mortality rate was higher in the first wave than in the second. We propose the following hypotheses that may contribute to the lower-case fatalities in the second wave. First, the decrease in case fatalities in the second wave compared to the first wave could be a harvesting effect. In other words, a large number of elderly people and people with comorbidities (the vulnerable groups) probably died in the first wave, especially in countries with high infection rates. Second, testing capacity, improved case management, and health system strengthening were better reinforced in the second wave. Third, in several countries, the age structure of those infected may have changed between the first and second waves for many reasons. For example, the social movement in many countries might involve more young and healthy individuals, but in our country, this was not necessarily found because we had a higher average age in the second wave than in the first ^19^. This improvement in the outcome of admitted patients could be related to the fact that the health care system in our country, as in many others, has become better prepared. We have more experience and better treatments. We do more diagnostic tests, which allows us to detect serious cases at an early stage and treat them more effectively. and receive more effective treatments. In this regard, in the second wave, patients were treated more frequently with dexamethasone, as suggested by the RECOVERY study. In addition, the number of health care providers was higher in the second wave compared to the first wave ^20^.

The predictors of mortality in both waves were respiratory distress and severe COVID-19 stage. In the first wave of over 60 years, respiratory distress, desaturation (<89%) and severe COVID-19 stage emerged as factors associated with death, while in the second wave it was mainly respiratory distress, desaturation (<89%) and severe stage. Age is no longer associated with mortality as clinicians have drawn particular attention to the elderly as previous studies in the first wave of DRC had already emphasized age as a mortality factor ^21-23^. Severe stage and desaturation still remain a great challenge in the management of COVID-19, especially when the patient consults late. As we observed, patients in the first wave came late, some with saturation below 50% in greater numbers than in the second wave.

### Limitations

Our study has some limitations. First, because of the retrospective study design, not all laboratory tests were performed on patients, including d-dimer, IL-6, troponin, lactate dehydrogenase, and serum ferritin. Therefore, their role may be underestimated in predicting in-hospital death. Second, missing data, especially in laboratory parameters, could lead to bias in the final analysis.

## Conclusion

The mean age was higher in the second wave. The essential therapeutic approaches in COVID-19, including corticosteroid therapy and anticoagulation, were more used in the second wave. Mortality was higher in the first wave. The factors predicting mortality that emerged in both waves were respiratory distress and severe stage of COVID-19, whereas age no longer emerged as a factor in mortality in the second wave. Health system improvement and education of those at high risk of mortality should be pursued to continue to reduce the mortality of patients hospitalized with COVID-19.

## Data Availability

Datasets used and analyzed during the current study are available from the corresponding author on request.

## Author contributions

Conceptualization: Madone Mandina and JR Makulo.

Methodology: Madone Mandina, JR Makulo, Ben Bepouka and Ossam Odio

Validation: Madone Mandina, JR Makulo, Ben Bepouka and Ossam Odio

Writing – Original Draft: Madone Mandina, Jean-Robert Makulo, Roger Wumba, Ben Bepouka, Jerome Odio, Aliocha Nkodila, Murielle Longokolo, Nadine Mayasi,Donatien Mangala, Guyguy Kamwiziku, Auguy Luzayadio Longo, Guillaume Mpia, Yamin Kokusa, Hervé Keke, Marcel Mbula, Hippolyte Situakibanza, Ernest Sumaili, Jean-Marie Kayembe

